# The Struggle to Vaccinate: Unveiling the Reality of the first year of Covid-19 Vaccination in the Democratic Republic of Congo

**DOI:** 10.1101/2024.01.03.24300795

**Authors:** Amani Adidja, Cikomola Mwana Bene Aimé, Christophe Lungoyo Luhata, Arsène Kabwaya Mukoka, Fabrice Zobel Lekeumo Cheuyem, Samuel Mpinganjira, Dumisile Sibongile Nkosi, Kimberly Cheryl Chido Konono, Michael Ngigi, Pierre Ongolo-Zogo

## Abstract

**Introduction:** The emergence of COVID-19 as a global pandemic has affected countries worldwide, including the Democratic Republic of Congo (DRC). The DRC has experienced four waves of COVID-19, each associated with a new variant of the virus. To control the spread of the virus, the government of the DRC implemented various measures, including vaccination. The country developed a COVID-19 vaccine deployment plan, targeting high-risk groups, and launched a vaccination campaign in April 2021. This study aims to comprehensively assess the COVID-19 vaccination program in the DRC during its first year of implementation, including progress, coverage, types of vaccines administered, and a comparison with other neighboring countries.

**Methods:** This study was an analysis of the COVID-19 vaccination data during its first year of implementation in DRC. Data were collected from multiple sources, including the Ministry of Health and the WHO, and analyzed using descriptive statistics. The study received clearance and used de-identified and aggregated data.

**Results:** Out of the 26 provinces in the country, only 15 began immunization activities with varying levels of coverage, ranging from 0.02% to 6.91%. The number of functional vaccination sites remained patchy across the country. By March 2022, 5.7% of the population had received at least one vaccine dose, with 1.03% fully vaccinated. In most provinces, men were more compliant with vaccination than women. More than half of the vaccinated individuals preferred the Janssen vaccine. Compared to neighboring countries, the DRC has lagged behind in its vaccination efforts, having administered only 1.1 million of doses received (8%) and has vaccinated only 2% of its population with at least one dose, the lowest among the countries analyzed.

**Conclusion:** Despite the challenges faced in the first year of the COVID-19 vaccination, DRC has made significant progress in vaccinating its population. The slow progress highlights the need for continued investment in health systems. These insights can inform future Covid-19 vaccination campaigns in DRC and other low-income countries.

## Introduction

Coronaviruses are zoonotic infections that can be transmitted from animals to humans and from one person to another [1]. They can cause various diseases in humans, ranging from the common cold to severe acute respiratory syndrome (SARS) [2]. In December 2019, a novel coronavirus called SARS-CoV-2 was identified as the cause of a new infectious respiratory disease in the city of Wuhan in China and was later named COVID-19 [3]. COVID-19 then spread worldwide and was declared a pandemic by the World Health Organization (WHO) on 11 March 2020 [4].

Many vaccines was developed and approved by the WHO and are currently being used worldwide to control the spread of COVID-19 [12]. COVAX was the vaccines pillar of the Access to COVID-19 Tools (ACT) Accelerator, a global initiative that was established in April 2020, for the production, and equitable access to COVID-19 tests, treatments, and vaccines [13]. It was co-convened by the Coalition for Epidemic Preparedness Innovations, the Vaccine Alliance (Gavi) and the WHO working in partnership with United Nation Children’s Fund (UNICEF) as a key implementing partner [14]. The objective of COVAX was to ensure rapid, fair, and equitable access to COVID-19 vaccines worldwide and enable countries to have access to the world’s largest and most diverse COVID-19 vaccine portfolio [13,15]. As of April 2022, out of the 47 countries in the WHO-Africa Region, 17.1% had completed their primary series, compared to the global rate of 60% [16].

In February 2020, Covid-19 was first reported in Africa when Egypt notified its first case [5]. Other African countries subsequently confirmed their first cases including the Democratic Republic of the Congo (DRC) on March 10^th^, 2020 [6]. The situation in the DRC rapidly worsened, with the virus spreading to multiple provinces, such as Kinshasa, Kwilu, Ituri, North Kivu, South Kivu, Haut Katanga, and Kongo Central [7]. The government of the DRC therefore declared a state of emergency on March 24^th^, 2020 [8]. The country has experienced four waves, the first two of which were the most severe, and each waves was associated with a new variant of the SARS-CoV-2 virus [9,10]. The address this health crisis, several measures including vaccination were implemented to curb the outbreak in the country [11].

In January 2021, the Democratic Republic of Congo (DRC) developed a Covid-19 Vaccine deployment plan in line with the recommendations of the national technical advisory group for vaccines and the WHO guidelines [17]. The plan prioritized high-risk groups such as healthcare workers, people living with co-morbidities and those aged 55 and over, with a total target of 53,984,184 people aged 18 and over. In April 2021, the Ministry of Health launched a Covid-19 vaccination campaign, with the aim of vaccinating at least 20% of the population by the end of 2021. With the introduction of vaccination programme in the country, it was crucial to monitor and review progress after one year of implementation. Understanding the COVID-19 vaccination program in the first year of implementation is critical for both health system planning, preparedness and improving future uptake of vaccines [18]. The implementation of Covid-19 vaccination program in its first year has not been well described in previous studies. Research conducted where either limited in scope like addressing one province [19,20] or one component of the vaccination for instance, the communication, the demand generation [21–24]. The only study that gave an overview of implementation was a commentary but was limited as it only focused on ChAdOx1[25]. While commentaries can provide valuable insights and perspectives, they are generally considered inferior to a full research article because they lack the same level of rigor and systematic methodology as empirical research [26]. To the best of our knowledge, this study is the first that assess the implementation progress during the early stage of implementation that gives an appreciation and perspective to understand if the situation was similar across all the Central Africa countries. We therefore aimed to fill this gap, by performing a review with the objective to provide insights on the first-year of the Covid-19 vaccination in DRC.

## Methods

### Study design

This was a secondary analysis of the COVID-19 vaccination programme in the Democratic Republic of Congo (DRC) in the first year of implementation, from April 2021 to March 2022 [27].

### Setting

The Democratic Republic of Congo (DRC) is the largest country in Sub-Saharan Africa, covering a surface area of 1,027 km² and situated in Central Africa [28]. The country borders nine other countries and is divided into 26 provinces, namely: Bas-Uele (Lower Uele), Equateur, Haut-Katanga (Upper Katanga), Haut-Lomami (Upper Lomami), Haut-Uele (Upper Uele), Ituri, Kasai, Kasai-Central, Kasai-Oriental (East Kasai), Kinshasa, Kongo Central, Kwango, Kwilu, Lomami, Lualaba, Mai-Ndombe, Maniema, Mongala, Nord-Kivu (North Kivu), Nord-Ubangi (North Ubangi), Sankuru, Sud-Kivu (South Kivu), Sud-Ubangi (South Ubangi), Tanganyika, Tshopo, Tshuapa;. The DRC has a complex health system with three levels of organization: central, intermediate, and operational. At the operational level, there are 519 Health Zones, 393 General Referral Hospitals, and 8,504 health areas [29]. The population is estimated of 108 million, with a median age of 16.7 years and a population growth rate of 3.1% [28]. The country is one of the poorest in the world, with over 70% of the population living in poverty and 63% living below the poverty line [30]. The majority of the population is Christian, with over 93% identifying as such, of which 42% are Catholics [31]. The country has a long history of armed civil unrest, resulting in a complex humanitarian emergency (CHE) and a public health context that is plagued by recurrent disease outbreaks as yellow fever, polio, cholera, measles, and Ebola. with the highest count of public health emergencies (PHEs) in Africa, with an estimated 5.5 million internally displaced persons (IDPs) and 526,370 refugees and asylum-seekers, mainly from Burundi, the Central African Republic, and South Sudan [32–34].

### Data collection

Data were collected from multiple sources, including the Ministry of Health of the DRC and the WHO-African Region Covid-19 vaccination database. The Ministry of Health also collects monthly data on programmatic indicators, such as vaccination strategies and vaccine procurement. These data are officially reported by provinces based on a standard reporting protocol, and the reporting timeframe is comparable across all provinces. Data collection tools including Population-based immunizations registers have been made available to providers to capture information on the vaccine administered.

### Variables

A paper-based tool was used to collect variables related to the COVID-19 vaccination implementation. These variables include age, gender, health areas, population, vaccination start date, health workers’ status, doses administered, first and second doses, vaccine management, at least one health condition, date of administration, vaccine type, doses based on chronological order. All data were recorded on paper using various registers and forms, including vaccination registers, a registry of stocks, and vaccination cards.

### Data analysis

All the statistical analysis and visualization of study data were done using the Microsoft Office Excel 2016. We summarized using descriptive statistics such as the for numerical continuous variables, counts and proportions. The results were presented in tables, line graphs and maps. Regarding data related to vaccination progress in neighboring African countries, the estimated doses needed to vaccinate 70% of the population were presented alongside the doses received, and the percentage of doses received over the amount needed to reach 70% of the population fully vaccinated.

## Results

### COVID-19 Vaccination Coverage across Provinces

Out 26 provinces of the country, 16 started immunization activities in the first year of implementation. The highest coverage of fully vaccinated individual was observed in Kasai Oriental province (6,91 %) followed by Haut Uele (2,51 %) and Sud – Ubangi (2,85 %). On the other hand, Kwilu (0,07 %), Tshuapa (0,03 %) and Equateur (0,02 %) presented the lowest performance (Fig 1).

**Fig 1.**
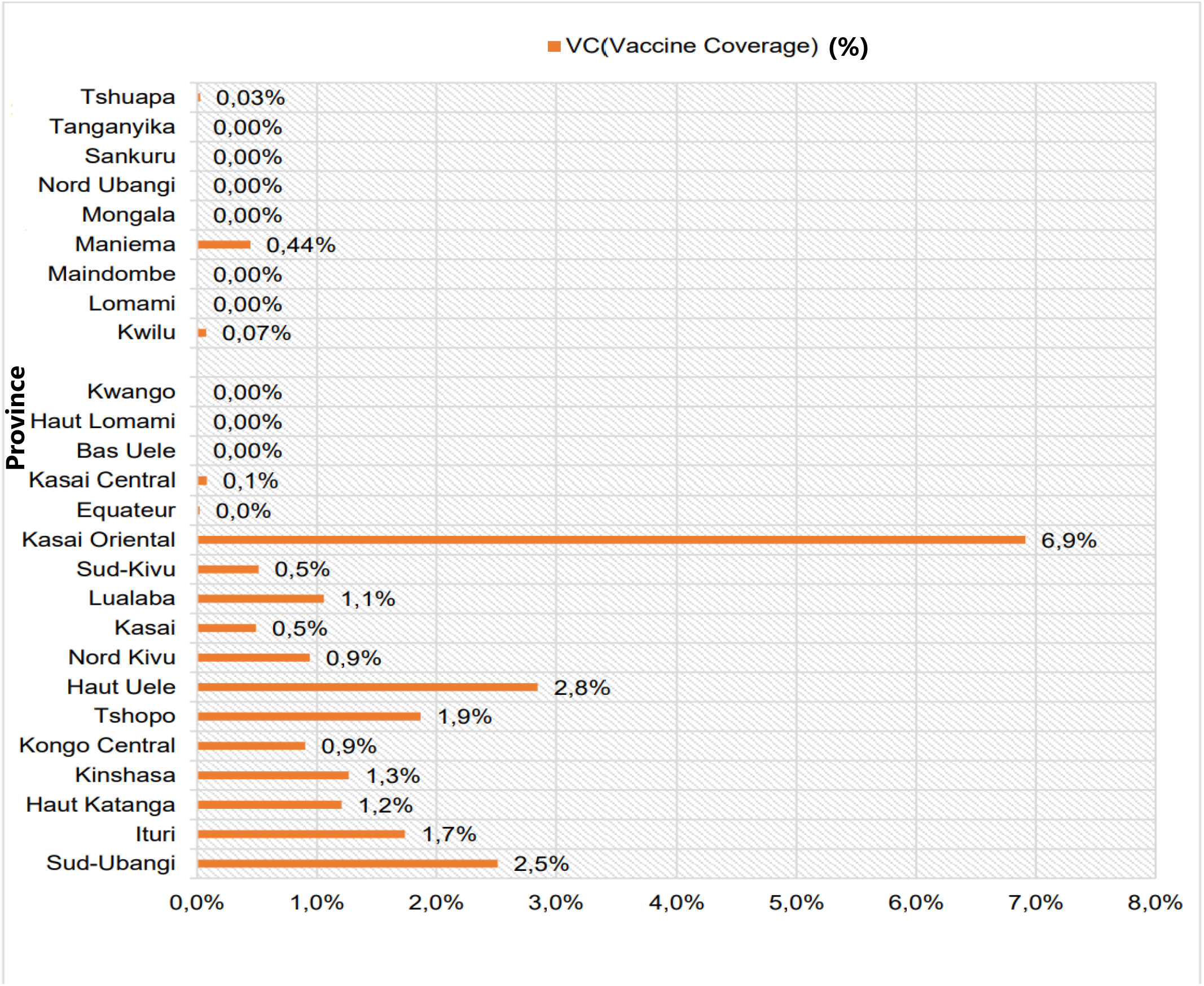
Coverage of fully vaccinated people by province from May 2021 to April 2022 in DRC.

### Functional Vaccination Sites in the Country

Data suggest that vaccination efforts in the DRC have improved over time, with a gradual increase in the number of functional vaccination sites from May 2021 to April 2022. There was a considerable variation in the number of functional vaccination sites across with a patchy distribution across different provinces. Kinshasa had the highest number of sites in all months, while some provinces like Kasaï and Tshuapa had only a few sites, or even none in some months. Haut-Katanga had a steady increase in the number of sites from May to October 2021, but a significant decrease in November and a slight increase in December. In contrast, Nord-Kivu had a fluctuating number of sites, with a peak in February 2022. Kinshasa had a consistently high number of sites throughout the entire period, while other provinces like Kasaï central, kwilu, and Maniema had very few sites, or none at all, in most months (Table 1).

**Table 1.** Functional COVID-19 vaccination sites in the first year of implementation from April 2021 to April 2022 in DRC.

| Province | 2021 |  |  |  |  |  |  |  |  | 2022 |  |  |  |
| --- | --- | --- | --- | --- | --- | --- | --- | --- | --- | --- | --- | --- | --- |
|  | Apr | May | June | Jul | Aug | Sept | Oct | Nov | Dec | Jan | Feb | Mar | Apr |
| Bas-Uele | 0 | 0 | 0 | 440 | 0 | 0 | 0 | 0 | 0 | 0 | 0 | 0 | 1157 |
| Équateur | 0 | 0 | 0 | 0 | 440 | 726 | 726 | 726 | 726 | 726 | 726 | 726 | 726 |
| Haut-Katanga | 0 | 3222 | 5689 | 12154 | 13129 | 18241 | 20716 | 28770 | 33394 | 50552 | 55700 | 62629 | 66474 |
| Haut-Lomami | 0 | 0 | 0 | 0 | - | 0 | 0 | 0 | 0 | 0 | 0 | 0 | 0 |
| Haut-Uele | 0 | 0 | 0 | 3771 | 3771 | 5697 | 6040 | 9128 | 9262 | 12900 | 35049 | 39308 | 39308 |
| Ituri | 0 | 0 | 0 | 4049 | 6096 | 8574 | 9164 | 9164 | 13827 | 48237 | 48237 | 83399 | 83399 |
| Kasai | 0 | 0 | 0 | 0 | - | 0 | - | 0 | 0 | 13804 | 13804 | 13804 | 181641 |
| Kasai central | 0 | 0 | 0 | 0 | 0 | 278 | 278 | 278 | 278 | 590 | 2364 | 5205 | 5560 |
| Kasai oriental | 0 | 0 | 0 | 733 | 733 | 1029 | 1221 | 1221 | 1221 | 4688 | 255403 | 295830 | 295830 |
| Kinshasa | 4165 | 4773 | 7593 | 28682 | 31417 | 41356 | 57499 | 78939 | 118126 | 131629 | 136545 | 176374 | 315662 |
| Kongo Central | 0 | 0 | 0 | 10990 | 10696 | 15814 | 16546 | 16546 | 17127 | 29804 | 31612 | 32905 | 33589 |
| Kwango | 0 | 0 | 0 | 0 | 0 | 0 | 0 | 0 | 0 | 0 | 0 | 0 | 0 |
| Kwilu | 0 | 0 | 0 | 192 | 192 | 192 | 192 | 192 | 356 | 192 | 192 | 2595 | 70688 |
| Lomami | 0 | 0 | 0 | 0 | 0 | 0 | - | 0 | 0 | 0 | 0 | 0 | 0 |
| Lualaba | 0 | 0 | 0 | 6488 | 6488 | 8445 | 8733 | 11637 | 12181 | 16185 | 27949 | 27952 | 27952 |
| Mai-Ndombe | 0 | 0 | 0 | 0 | 0 | 0 | 0 | 0 | 0 | 0 | 0 | 0 | 0 |
| Maniema | 0 | 0 | 0 | 0 | 0 | 0 | 0 | 0 | 0 | 0 | 130 | 6682 | 6682 |
| Mongala | 0 | 0 | 0 | 0 | 0 | 0 | 0 | 0 | 0 | 0 | 0 | 0 | 0 |
| Nord-Kivu | 0 | 1499 | 6789 | 6954 | 7882 | 10283 | 13064 | 18903 | 25017 | 65661 | 71969 | 82836 | 89471 |
| Nord Ubangi | 0 | 0 | 0 | 0 | 0 | 0 | 0 | 0 | 0 | 0 | 0 | 0 | 0 |
| Sankuru | 0 | 0 | 0 | 0 | 0 | 0 | 0 | 0 | 0 | 0 | 0 | - | 0 |
| Sud-Kivu | 0 | 493 | 3786 | 3896 | 4092 | 5545 | 6022 | 7013 | 9030 | 17108 | 37397 | 38933 | 38933 |
| Sud-Ubangi | 0 | 0 | 0 | 565 | 565 | 821 | 941 | 4018 | 10348 | 31054 | 47472 | 53717 | 53717 |
| Tanganyika | 0 | 0 | 0 | 0 | 0 | 0 | 0 | 0 | 0 | 0 | 0 | 0 | 0 |
| Tshopo | 0 | 0 | 0 | 323 | 323 | 646 | 547 | 547 | 3335 | 26267 | 51121 | 70754 | 70754 |
| Tshuapa | 0 | 0 | 0 | 0 | 0 | 0 | 0 | 0 | 0 | 0 | 0 | 526 | 526 |
| **Total** | **4165** | **9987** | **23857** | **79237** | **85824** | **117647** | **141689** | **187082** | **254228** | **449397** | **815670** | **994175** | **1382069** |

### Trends of COVID-19 Vaccination Coverage

As of March 2022, around 5.7% of the population had received at least one dose of the vaccine, and 1.03% were fully vaccinated. The initial four months of the vaccine rollout experienced a low monthly uptake for the first dose, indicating a lackluster vaccination drive. However, February witnessed a surprising turn of events as the number of vaccine doses administered spiked, reaching a staggering 378,782, which was the highest uptake recorded. Regrettably, the positive momentum was not sustained, as the number of vaccine doses administered decreased progressively after that peak. Moreover, no significant changes were observed in the uptake of the second vaccine dose during the same period (Fig 2).

**Fig 1.**
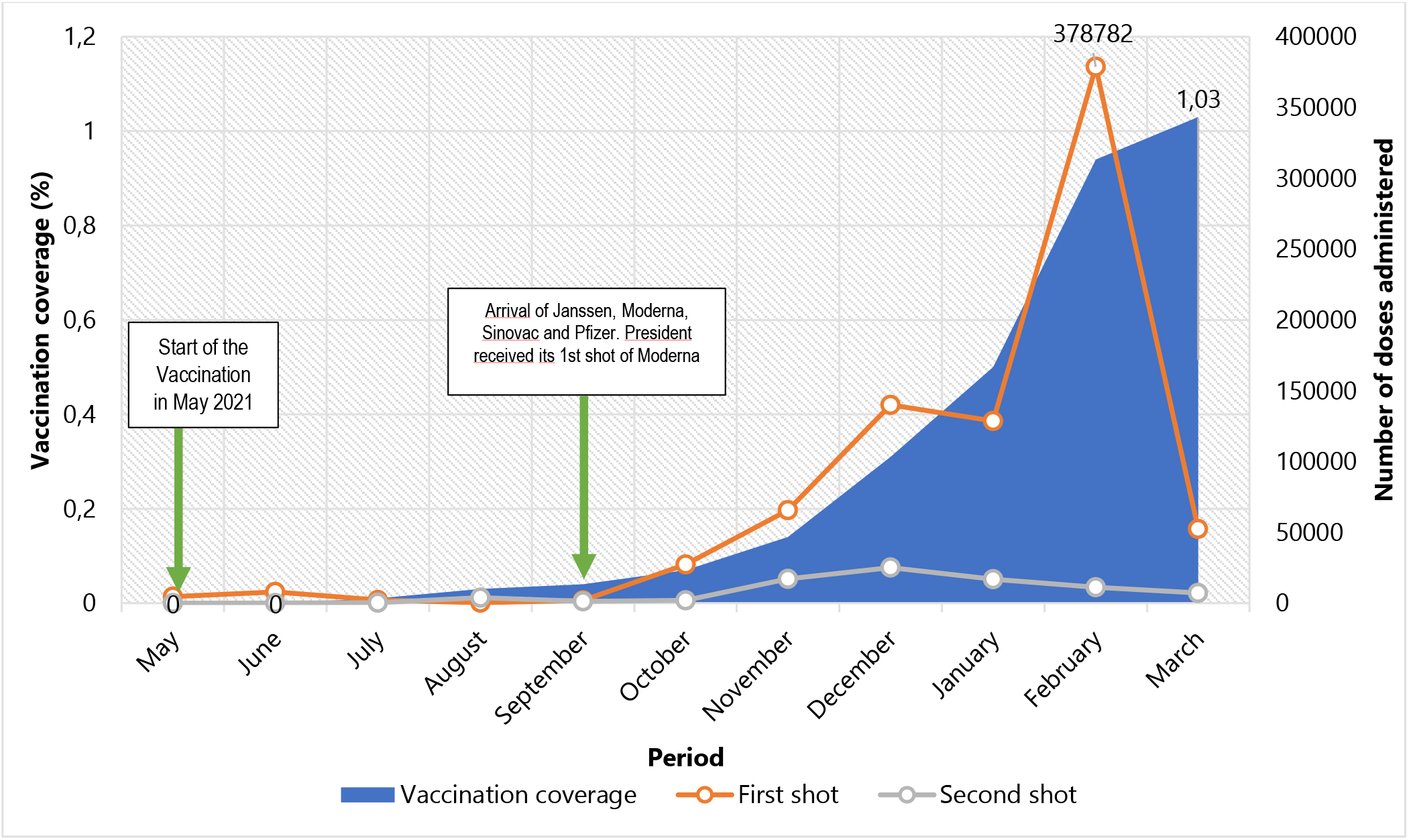
Trends in coverage of COVID-19 vaccination during the first year of implementation in DRC, April 2022.

### Gender Comparison among Vaccinated Population

with the exception of the Kasai Oriental province, the total number of men vaccinated against COVID-19 in each province of the Democratic Republic of the Congo (DRC) was higher than the number of vaccinated women (Fig 3).

**Fig 2.**
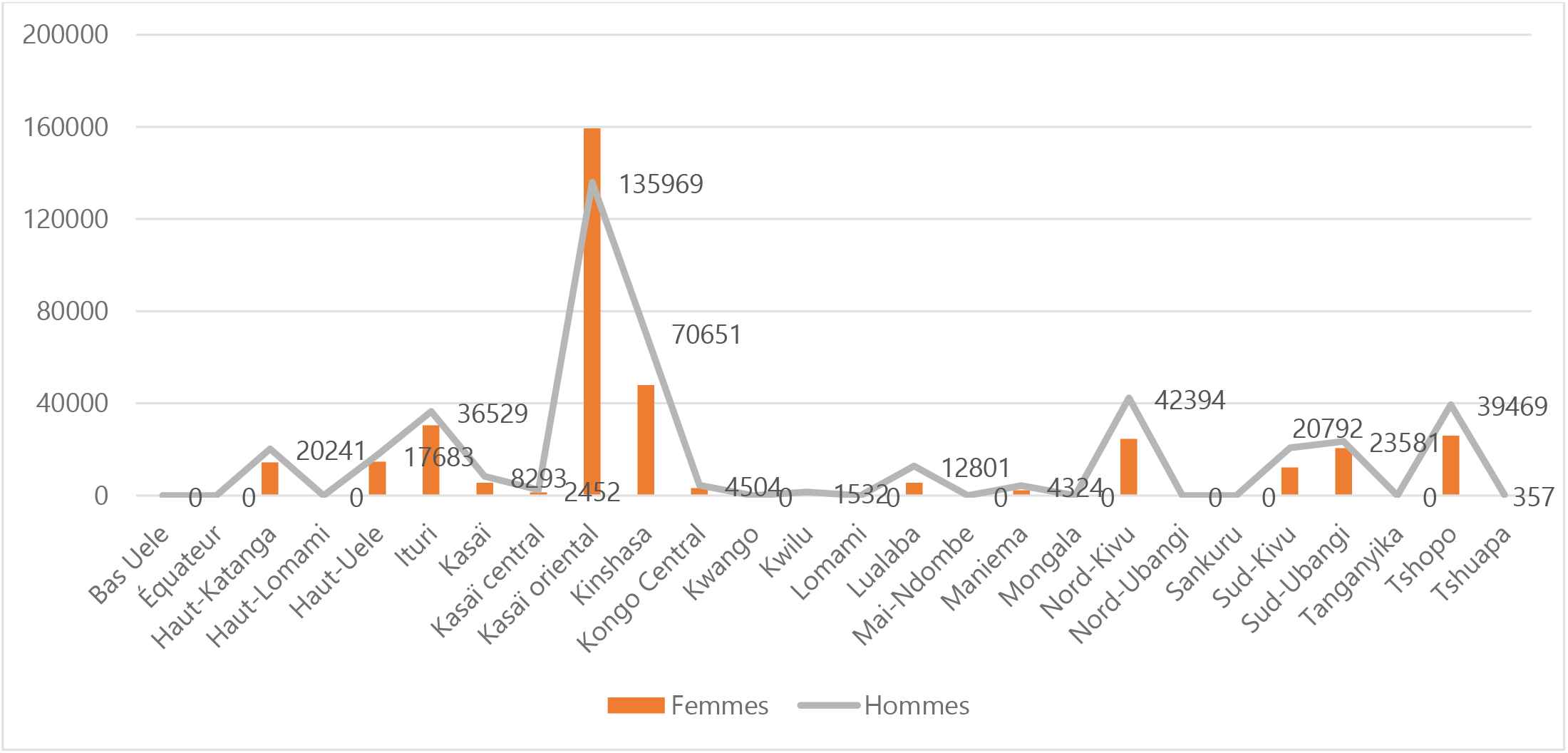
Vaccinated people according to the gender and province of residence in the first year of COVID-19 immunization activities in DRC, April 2022.

### Mapping of High Risks Groups Vaccinated

Overall, the data indicates that there are 8,302 Healthcare Workers (HW) and 46,495 elderly individuals that have completed their primary vaccine series. The data reveals that Kinshasa has the highest number of individuals who have completed their primary vaccination series, with 2,182 HWs and 10,813 elderly individuals, followed by Nord-Kivu with 2,546 HWs and 9,803 older persons, and Sud-Kivu with 776 HWs and 10,219 elderly individuals. On the other hand, Kasaï had the lowest number of individuals who have completed their primary vaccination series, with only 14 elderly individuals. Furthermore, the data indicates that a significant number of vaccinated people with at least one dose living with comorbidities are present in Haut-Katanga (n=3,385) followed by Kasaï oriental (n=2,152) and Haut-Uele(1,031 individuals) (Fig 4,5 and 6)

**Fig 4.**
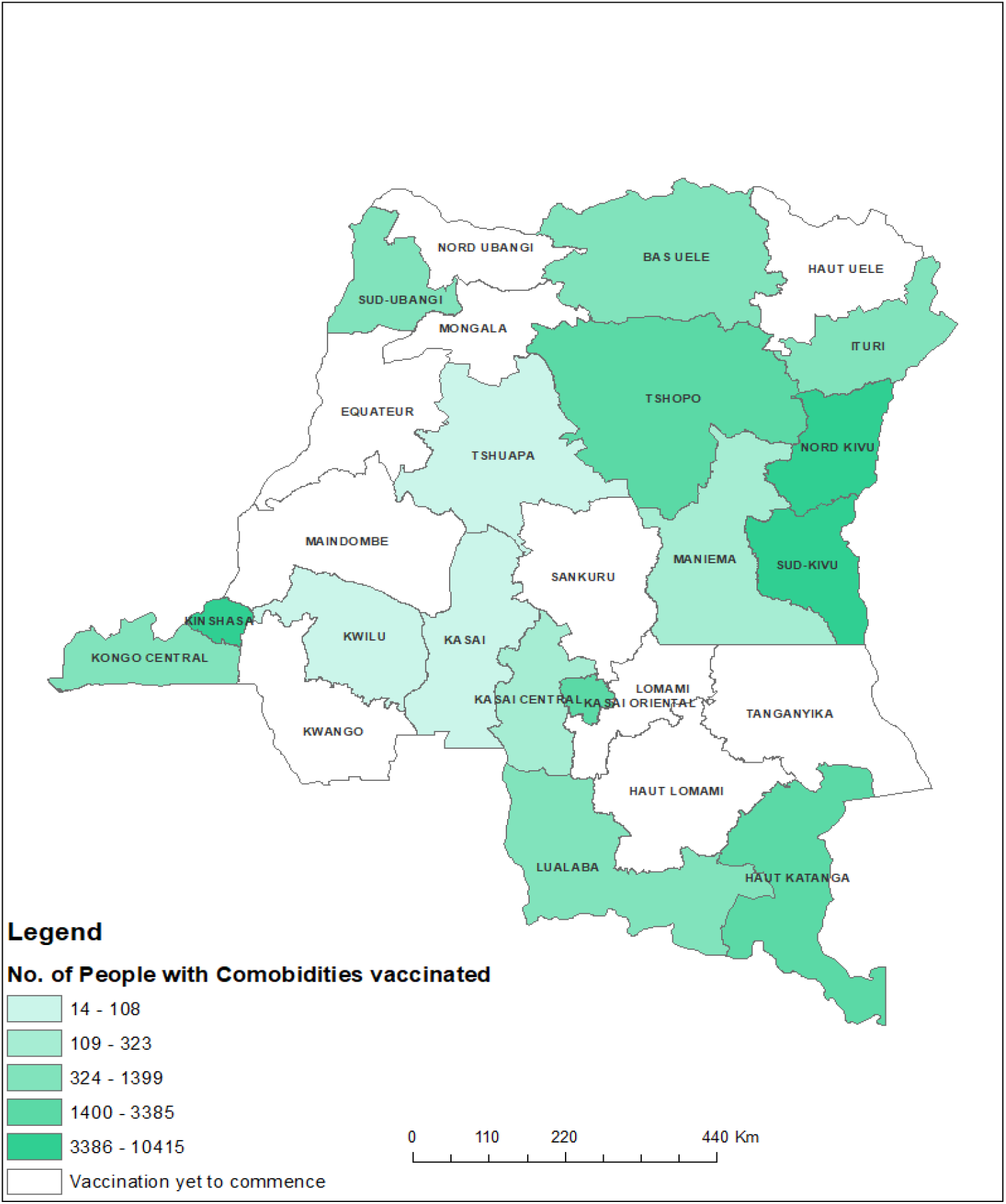
Number of people living with comorbidities vaccinated between May to Arpil 2022 in DRC.

**Fig 5.**
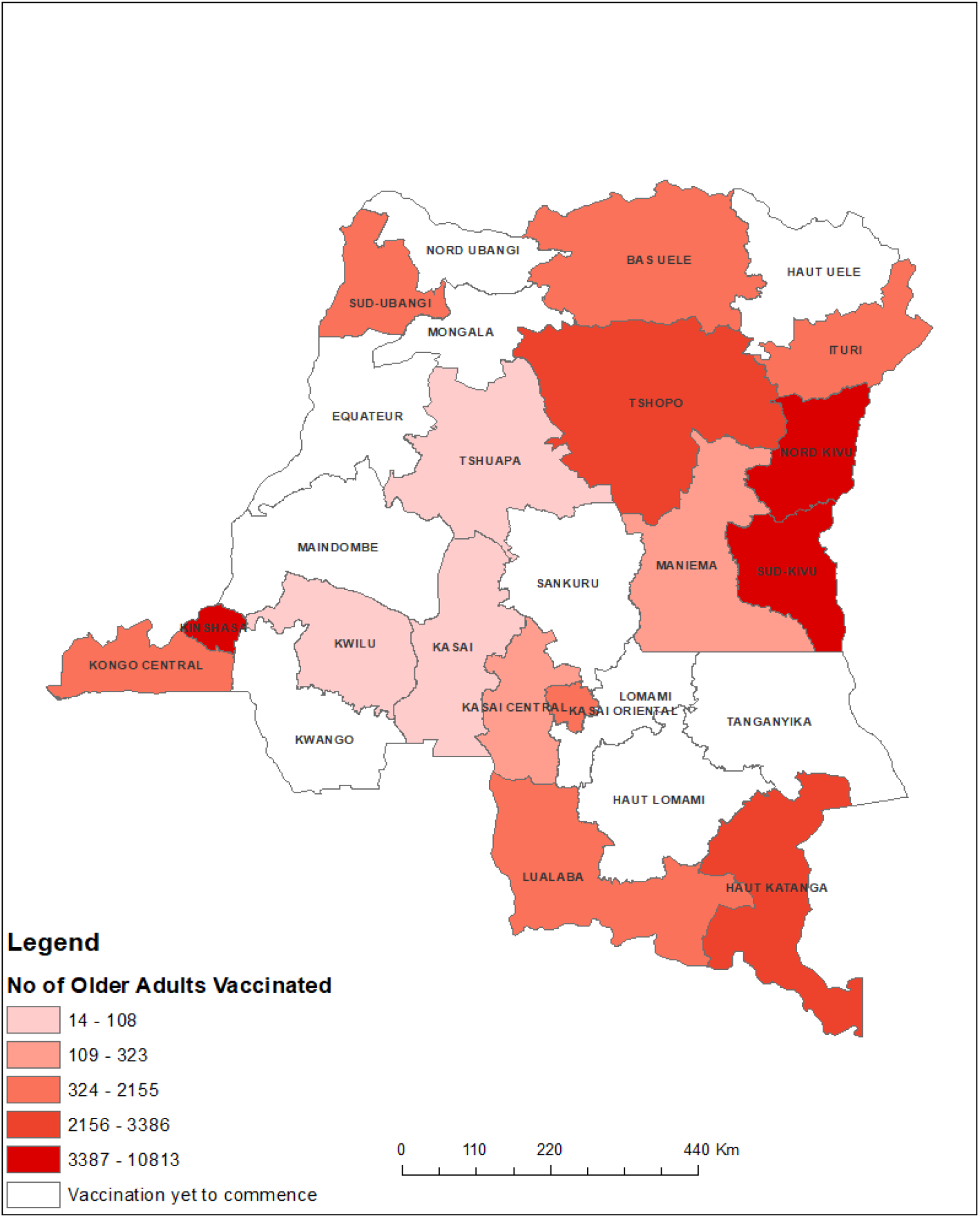
Number of vaccinated older adults between May to Arpil 2022 in DRC.

**Fig 6.**
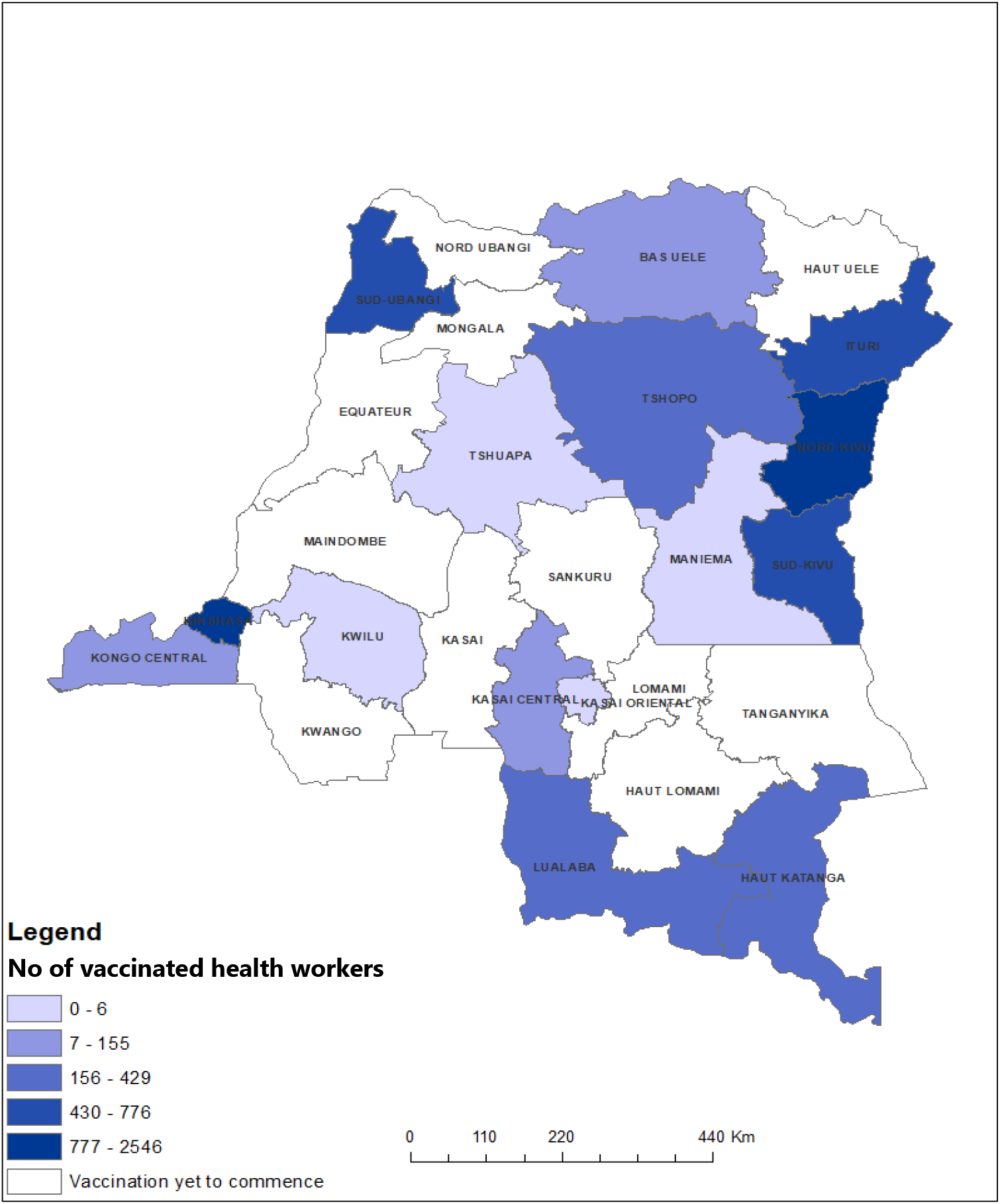
Number of vaccinated older adults between May to Arpil 2022 in DRC.

### Type of Vaccines Administered

The country used a range of vaccines, including the AstraZeneca, Johnson & Johnson, Pfizer and Sinovac vaccines. A little more than half of vaccinated individual prefer the Janssen vaccine (48%) followed by Moderna (25%) and Pfizer (17%), AstraZeneca was the less requested among available COVID-19 vaccine in the country (2%) (Fig 7).

**Fig 7.**
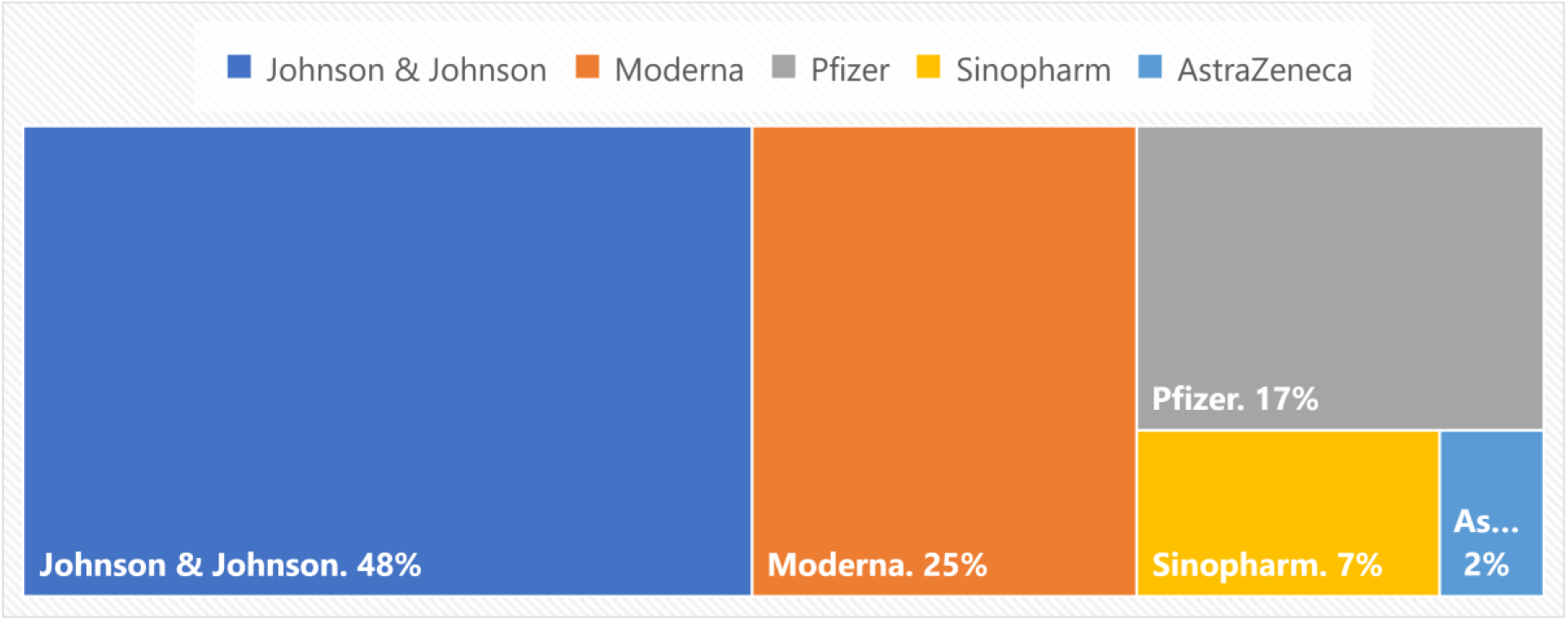
Types of vaccines administered from April 2021 of March 2022 in DRC.

### Comparison with Vaccination Progress in Neighboring Countries

Angola had the highest percentage of doses received over the estimated doses needed to vaccinate 70% of its population, with 84.43%. In contrast, Cameroon had the lowest percentage of doses received, with only 9%. Sao Tome and Principe had the highest percentage of the population that had received at least one dose, with 51.90%, while DRC had the lowest percentage with only 2%.

Central African Republic had the highest percentage of doses administered over the doses received at 40%, while the Democratic Republic of the Congo (DRC) had the lowest percentage at 8%. DRC received 14,394,340 doses of the COVID-19 vaccine, which is 11.48% of the estimated doses needed to vaccinate 70% of its population. In contrast, other countries such as Angola, Equatorial Guinea, and Gabon have received a higher percentage of the estimated doses needed to vaccinate 70% of their population. The DRC had administered 1,143,186 doses of the vaccine, which was only 8% of the doses received. Additionally, the DRC had vaccinated only 2% of its population with at least one dose, which was the lowest among the countries in the table. Furthermore, the DRC administered 1.1 million doses, which was 8% of the doses received. This proportion was also lower than other countries, such as Equatorial Guinea, which administered 57% of the doses received, and the Central African Republic, which administered 40% of the doses received.

The proportion of DRC’s population that received at least one dose of the vaccine was 2%, which is the lowest among all the countries in the table. In contrast, Sao Tome and Principe have vaccinated 49.7% of their population with at least one dose of the vaccine.

**Table 2.** COVID-19 vaccine rollout in the 10 Central African countries from April 2021 until March 2022 in DRC.

| Country | Vaccine dose |  |  | Target<br>*n* | Vaccination coverage |  |  |
| --- | --- | --- | --- | --- | --- | --- | --- |
|  | Needed | Received | Administered/<br>Received |  | Received<br>dose 1 | CPS | Total doses<br>administered/<br>Target (%) |
|  | *n* | *n* (%) | *n* (%) |  | (%) | *n* (%) |  |
| Angola | 46012775 | 38848267 (84.4) | 17896626 (46) | 12059919 | 36.7 | 6327907 (19.3) | 54.5 |
| Cameroon | 37164210 | 3344550 (9.0) | 1821134 (54) | 1475516 | 5.6 | 1113944 (4.2) | 6.9 |
| CAR | 6761670 | 2568280 (37.9) | 1037580 (40) | 1015918 | 21 | 988591 (20.5) | 21.5 |
| Chad | 22996203 | 4485510 (19.5) | 2291152 (51) | 2212530 | 13.5 | 2087559 (12.7) | 13.9 |
| DRC | 125385966 | 14394340 (11.4) | 1143186 (8) | 1771324 | 2 | 1064851 (1.2) | 1.3 |
| Equatorial Guinea | 1964179 | 820000 (41.7) | 471006 (57) | 262703 | 18.7 | 206762 (14.7) | 33.6 |
| Gabon | 3116019 | 1630600 (52.3) | 556657 (34) | 300192 | 13.5 | 254453 (11.6) | 25 |
| Republic of Congo | 7725329 | 3236630 (41.9) | 830379 (26) | 693003 | 12.6 | 640464 (11.6) | 15 |
| Sao Tome and Principe | 306825 | 431020 (140.4) | 208657 (48) | 113708 | 51.9 | 86914 (39.7) | 95.2 |

CAR: Central African Republic; DRC: Democratic Republic of Congo; CPS: Complete Primary Series of COVID-19 vaccination

The data also reveals that DRC has a low rate of doses administered per 100 population (1.3), which is substantially lower than the other countries in the table. For example, the rate in Equatorial Guinea is 33.6 doses per 100 population, while Sao Tome and Principe has a rate of 95.2 doses per 100 population.

## Discussion

At the end of the first year of vaccination, only half of the provinces have started vaccination although the country launched its COVID-19 vaccination campaign on April 19, 2021[35]. Disparities between provinces may be explained by the slow rollout of COVID-19 vaccination in the first year, with only 57% of provinces reached by the end of the first year. By March 2022, over 5 million doses of COVID-19 vaccines had been administered in the country. The country began administering the Oxford-AstraZeneca vaccines manufactured by the Serum Institute of India in May 2021, starting with Kinshasa, Kongo Central, Haut Katanga, North Kivu, South Kivu, and Lualaba. As of July 10, 2021, vaccination efforts had been expanded to 344 vaccination sites in 13 provinces, reaching a total of 78,871 people with the first dose and 2,513 with the second [36]. By December 2021, only 0.87% of the population had received one dose of the COVID-19 vaccine [37].

The result in Kasai Oriental and the pic observed in March 2022 could be explained by the fact that in February 2022, a national coordination was established with a Steering Committee and Technical Committee, who developed a 3-month acceleration plan from April to June 2022 to intensify the implementation of the National Vaccine Deployment Plan (PNDV). This resulted in significant improvements, with over 2.5 million people vaccinated in three months, compared to only 800,000 in the previous 11 months according to the weekly Covid-19 National Taskforce report. Although progress has been made in immunizing its population, the country has fallen short of its national target of vaccinating 11 million people by the end of April 2022.

We performed a comparison of DRC with other nine countries in Central Africa, with the exception of Burundi who have been reluctant to vaccinate [38]. Despite this positive progress, the country has faced several challenges resulting in overall low immunization coverage, making it one of the lowest performing countries in the world for COVID-19 vaccination [39]. DRC has faced numerous challenges in its efforts to combat COVID-19 through vaccination, resulting in an overall low immunization coverage despite receiving 8.2 million doses of vaccines, only 528,000 were administered, representing just 11% of the available doses.

Low uptake may be attributed to the false start where the country refrained from vaccinating its population with the Oxford/AstraZeneca COVID-19 vaccines due to safety concerns in western countries, as well as adverse events associated with the AstraZeneca vaccine and the miscommunication surrounding it further exacerbating vaccine hesitancy and decreasing demand as such, the vaccination has been halted due to reported AEFI post AstraZeneca administration [40]. However, it resumed vaccination on August 17, 2021, after receiving a new batch of 51,840 Astra Zeneca doses. In September 2021, the country received three other types of vaccines, including 250,320 doses of Moderna’s mRNA 1273 vaccine, 250,380 doses of Pfizer’s Comirnaty vaccine, and 400,000 doses of Coronavac/Sinovac. Moderna’s vaccine was introduced into the vaccination program on September 13, 2021, following the vaccination of the President of the Republic. The takeoff of the vaccination that was observed in September 2021 could be explained, by the fact in addition to AstraZeneca vaccines, 4 additional vaccines were introduced including Pfizer, Moderna, Sinovac, and Johnson and Johnson in September 2021 [41]. the vaccination program and there was a strong advocacy where it was seen publicly that the President of the republic has taken the Moderna Vaccines.

It is worth noting that the DRC is a large and populous country with a high burden of disease, including ongoing outbreaks with an Ebola outbreak stroked in Beni in early October 2021[42], as the country was introducing the covid-19 vaccines in additional provinces. The introduction of a new health crisis, such as the Ebola outbreak, in the midst of an ongoing immunization program has likely diverted attention and resources from the EPI program. The country has also faced several outbreaks of cholera, meningitis, yellow fever, poliomyelitis, and measles [43]. These competing priorities have made it difficult to allocate resources and attention to the COVID-19 response: This is because the same actors and infrastructures are often used to respond to both crises, which can lead to a strain on already limited resources. The diversion of resources and attention may have contributed to delays in achieving set immunization targets and revealed major gaps in health systems. In fact, the shortage of healthcare professionals is a significant challenge, with only 0.28 physicians per 10,000 population, far below the goal of 22.8 professional health workers per 10,000 stipulated by WHO. Furthermore, the first year if implementation has been marked by a strike of health workers due to misuse of pandemic response funds have also led to demotivation and partial strikes which has decreases service delivery points [44].

The logistics of COVID-19 vaccines in DRC has been another hurdle with large amount of expired or near-expiry vaccine doses that were unable to be used or that receiving bilateral donations of no EUL COVID-19 vaccines [45]. Some cross-cutting challenges, including short-lived vaccine shelf life, the risk of vaccine expiry, moreover, the supply of vaccines was further complicated by the introduction of five new vaccines targeting an adult population that is not typically prioritized by the immunization program. Although COVID-19 vaccination has encountered difficulties in the DRC, routine immunization programs show a coverage for the third dose of the Pentavalent vaccine in 2020 of 57% [46].

Reaching high-priority groups in the DRC has proven to be a challenging feat. The COVID-19 vaccination rollout initially failed to target these groups, and the country lacked experience in implementing public health programs tailored to their needs. A myriad of obstacles, including knowledge gaps, poor service delivery, and limited access to health services, has hindered the country’s efforts to vaccinate its high-risk populations effectively. To overcome these challenges, the integration of COVID-19 vaccination with other primary healthcare interventions such as the distribution of insecticide-treated nets is crucial [47]. Additionally, community engagement, popularizing immunization, and bundling COVID-19 vaccines with other health interventions can help improve vaccine coverage rates [48,49]. Leveraging religious and community engagement can also help increase vaccine acceptance and uptake. Despite the challenges faced in the COVID-19 vaccination rollout, the DRC has an opportunity to address these limitations by implementing innovative and evidence-based approaches that prioritize its high-risk populations.

## Strengths and Limitations

While this study provides valuable insights into the COVID-19 vaccination program in the DRC, there are several limitations to consider. The study faced challenges in accurately counting the high priority group’s denominators, tracking individuals’ vaccination history, and analyzing already aggregated data. The findings are highly contextual and may not reflect the situation nationwide. To address these limitations, additional research that includes key informants and uses disaggregated data may be necessary. Moreover, the pandemic’s impact on routine health care utilization may lead to underestimation of comorbidities in existing health care records. The validity of the study’s conclusions may be affected by these limitations, and further analysis is needed to identify the specific factors contributing to the slow vaccine rollout in the DRC and other countries with similar challenges.

## Conclusion

The first year of the COVID-19 vaccination campaign in the DRC has been a mix of achievements and challenges. The country has made significant some progress in vaccinating its population, despite the many challenges. The slow progress in COVID-19 vaccination coverage in the DRC underscores the need for continued efforts to investing in health systems to improve overall health outcomes. The country has provided several lessons for the global community on how to effectively respond to global health emergencies. These insights will be valuable to other countries facing as they can learn from the DRC’s experience.

## Data Availability

All data produced in the present work are contained in the manuscript

## Declaration

### Authors’ Contribution

Conceptualization, A.A.; methodology, A.A., A.C.M.B. and F.Z.L.C; software, validation, A.A. and A.K.M.; data curation, formal analysis, A.A, A.K.M. and F.Z.L.C; writing—original draft preparation, A.A.; writing—review and editing, A.A, F.Z.L.C., D.S.N., S.M., M.N., C.L.L., K.C.C.K., and P.O.-Z.; visualization, A.A.; supervision, A.A. and P.O.-Z. All authors have read and agreed to the published version of the manuscript.

### Ethical Approval Statement

The protocol was approved by Institutional Review Board (IRB) of the DRC Ministry of Health and the ethical clearance: PEV/DIR/199/NZW/2022 of the 18^th^ April 2022 issued. All data have been first de-identified, robustly anonymized then aggregated primarily at the heath area with a secondary level of aggregation at the provincial level. Highly aggregated data are not expected to include risks of re-identification of individuals or disclosure of sensitive information.

### Availability of data and materials

Not applicable.

## Acknowledgements

Our gratitude goes to DRC Ministry of Health who accepted to share national data with the study investigator.

## Reference

1. Salata C, Calistri A, Parolin C, Palù G. Coronaviruses: a paradigm of new emerging zoonotic diseases. Pathog Dis. 2019 Dec 1;77(9):1–5.

2. McIntosh K, Perlman S. Coronaviruses, Including Severe Acute Respiratory Syndrome (SARS) and Middle East Respiratory Syndrome (MERS). Mand Douglas Bennetts Princ Pract Infect Dis. 2015;1928–1936.e2.

3. Yang P, Wang X. COVID-19: a new challenge for human beings. Cell Mol Immunol. 2020 May;17(5):555–7.

4. Cucinotta D, Vanelli M. WHO Declares COVID-19 a Pandemic. Acta Bio-Medica Atenei Parm. 2020 Mar 19;91(1):157–60.

5. Marzouk M, Elshaboury N, Abdel-Latif A, Azab S. Deep learning model for forecasting COVID-19 outbreak in Egypt. Process Saf Environ Prot Trans Inst Chem Eng Part B. 2021 Sep;153:363–75.

6. Nachega JB, Mbala-Kingebeni P, Otshudiema J, Zumla A, Tam-Fum JJM. The colliding epidemics of COVID-19, Ebola, and measles in the Democratic Republic of the Congo. Lancet Glob Health. 2020 Aug;8(8):e991–2.

7. Modeawi MN, Baya JL, Bosso B, Kobe JK, Kusagba JM, Magbukudua JM, et al. COVID-19 Pandemic in Democratic Republic of the Congo: An Opportunity for Economic Recovery. Br Int Exact Sci BIoEx J. 2021 May 5;3(2):103–13.

8. Nkashama SKK. Etat d’urgence sanitaire pour faire face à l’épidémie de covid-19 en République Démocratique du Congo. KAS Afr Law Study Libr. 2021;8(1):44–53.

9. Otshudiema JO, Folefack GLT, Nsio JM, Mbala-Kingebeni P, Kakema CH, Kosianza JB, et al. Epidemiological Comparison of Four COVID-19 Waves in the Democratic Republic of the Congo, March 2020-January 2022. J Epidemiol Glob Health. 2022 Sep;12(3):316–27.

10. Salyer SJ, Maeda J, Sembuche S, Kebede Y, Tshangela A, Moussif M, et al. The first and second waves of the COVID-19 pandemic in Africa: a cross-sectional study. Lancet Lond Engl. 2021 Apr 3;397(10281):1265–75.

11. Geneva, WHO. The World Health Assembly (WHA) 73 [Internet]. IFMSA. 2020 [cited 2023 Oct 30]. Available from: https://ifmsa.org/the-world-health-assembly-wha-73/

12. Forchette L, Sebastian W, Liu T. A Comprehensive Review of COVID-19 Virology, Vaccines, Variants, and Therapeutics. Curr Med Sci. 2021 Dec;41(6):1037–51.

13. Eccleston-Turner M, Upton H. International Collaboration to Ensure Equitable Access to Vaccines for COVID-19: The ACT-Accelerator and the COVAX Facility. Milbank Q. 2021 Jun;99(2):426–49.

14. Storeng KT, de Bengy Puyvallée A, Stein F. COVAX and the rise of the ‘super public private partnership’ for global health. Glob Public Health. 2021 Oct 22;1–17.

15. Md Khairi LNH, Fahrni ML, Lazzarino AI. The Race for Global Equitable Access to COVID-19 Vaccines. Vaccines. 2022 Aug;10(8):1306.

16. COVID-19 vaccination in the WHO African Region - 05 April 2022 [Internet]. WHO | Regional Office for Africa. 2023 [cited 2023 Oct 30]. Available from: https://www.afro.who.int/publications/covid-19-vaccination-who-african-region-05-april-2022

17. Guidance on developing a national deployment and vaccination plan for COVID-19 vaccines [Internet]. [cited 2023 Oct 30]. Available from: https://www.who.int/publications-detail-redirect/WHO-2019-nCoV-Vaccine-deployment-2021.1-eng

18. Amani A, Djossaya D, Njoh AA, Fouda AAB, Ndoula S, Abba-kabir HM, et al. The first 30 days of COVID-19 vaccination in Cameroon: achievements, challenges, and lessons learned. Pan Afr Med J [Internet]. 2022 Mar 14 [cited 2023 Oct 30];41(201). Available from: https://www.panafrican-med-journal.com/content/article/41/201/full

19. Prudence MN, Tsongo EM, Mbeva JBK, Edmond NN, Bonane JK, Guy MN, et al. Seroprevalence of anti-SARS-CoV-2 antibodies among hospital staff in North Kivu province, Democratic Republic of Congo. Int J Innov Appl Stud. 2022 Apr 2;36(1):67–77.

20. Mandefu SO, Emakanya BME, Kiangano EA, Tomilalo FO, Mandefu FO, Tukutuku ML, et al. Mapping of Routine Coronavirus Vaccination 2019 in Tshopo Province, DR Congo, July to December 2021. J Adv Med Pharm Sci. 2022 Nov 5;25–34.

21. Barrall AL, Hoff NA, Nkamba DM, Musene K, Ida N, Bratcher A, et al. Hesitancy to receive the novel coronavirus vaccine and potential influences on vaccination among a cohort of healthcare workers in the Democratic Republic of the Congo. Vaccine. 2022 Aug 12;40(34):4998–5009.

22. Ditekemena JD, Nkamba DM, Mutwadi A, Mavoko HM, Siewe Fodjo JN, Luhata C, et al. COVID-19 Vaccine Acceptance in the Democratic Republic of Congo: A Cross-Sectional Survey. Vaccines. 2021 Feb 14;9(2):153.

23. Mabasi Mayala G, Malonga Niangi L, Wembo Lombela G, Ntumba Kayembe JM. Première année de la pandémie à COVID-19 en République Démocratique du Congo: Revue de la gestion d’une crise dans un système de santé décentralisé. Ann Afr Méd En Ligne. 2022;4561–76.

24. Natuhoyila Nkodila A, Ngwala Lukanu P, Nlombi Mbendi C, Marie Tebeu P, Saint Antaon Saba J, Alex Kabangi Tukadila H, et al. Perception of the Congolese population on Covid-19 vaccination: cross-sectional survey of online. Int J Vaccines Vaccin. 2021;6(1):12–9.

25. Zola Matuvanga T, Doshi RH, Muya A, Cikomola A, Milabyo A, Nasaka P, et al. Challenges to COVID-19 vaccine introduction in the Democratic Republic of the Congo – a commentary. Hum Vaccines Immunother. 2022 Nov 30;18(6):2127272.

26. Sarma SK. Qualitative Research:Examining the Misconceptions. South Asian J Manag. 2015 Sep 1;22(3):176–91.

27. Sage Publications. SAGE research methods. Thousand Oaks, CA: SAGE Publications; 2011.

28. Congo, Democratic Republic of the. In: The World Factbook [Internet]. Central Intelligence Agency; 2023 [cited 2023 Oct 30]. Available from: https://www.cia.gov/the-world-factbook/countries/congo-democratic-republic-of-the/

29. République démocratique du Congo - Plan national de développement sanitaire recadré pour la période 2019-2022 : Vers la Couverture sanitaire universelle. [Internet]. [cited 2023 Oct 30]. Available from: https://www.ilo.org/dyn/natlex/natlex4.detail?p_isn=111796&p_lang=fr

30. Overview [Internet]. World Bank. [cited 2023 Oct 30]. Available from: https://www.worldbank.org/en/country/drc/overview

31. Priest RJ, Ngolo A, Stabell T. Christian Pastors and Alleged Child Witches in Kinshasa, DRC. OKH J Anthropol Ethnogr Anal Eyes Christ Faith [Internet]. 2020 Jan 30 [cited 2023 Oct 30];4(1). Available from: https://okhjournal.org/index.php/okhj/article/view/81

32. Social science support for COVID-19: Lessons Learned Brief 3 - Humanitarian programme recommendations for COVID-19 based on social sciences evidence from the DRC Ebola outbreak response - World | ReliefWeb [Internet]. 2020 [cited 2023 Oct 30]. Available from: https://reliefweb.int/report/world/social-science-support-covid-19-lessons-learned-brief-3-humanitarian-programme

33. Global Humanitarian Overview 2022 | Global Humanitarian Overview [Internet]. [cited 2023 Oct 30]. Available from: http://gho-2022-site.docksal.site/

34. Vaccination in Humanitarian Emergencies: Implementation Guide [Internet]. [cited 2023 Oct 30]. Available from: https://www.who.int/publications-detail-redirect/WHO-IVB-17.13

35. La vaccination contre le coronavirus, COVID-19 a commencé en République Démocratique du Congo [Internet]. OMS | Bureau régional pour l’Afrique. 2023 [cited 2023 Oct 30]. Available from: https://www.afro.who.int/fr/news/la-vaccination-contre-le-coronavirus-covid-19-commence-en-republique-democratique-du-congo

36. Wise J. Covid-19: European countries suspend use of Oxford-AstraZeneca vaccine after reports of blood clots. BMJ. 2021 Mar 11;n699.

37. Covid-19 Vaccination in the WHO African Region Monthly bulletin [Internet]. WHO | Regional Office for Africa. 2023 [cited 2023 Oct 30]. Available from: https://www.afro.who.int/health-topics/coronavirus-covid-19/vaccines/monthly-bulletin

38. Learning from Burundi’s political pivot on COVID-19 vaccines [Internet]. 2022 [cited 2023 Nov 1]. Available from: https://blogs.worldbank.org/nasikiliza/learning-burundis-political-pivot-covid-19-vaccines

39. Impouma B, Mboussou F, Farham B, Makubalo L, Mwinga K, Onyango A, et al. COVID-19 vaccination rollout in the World Health Organization African region: status at end June 2022 and way forward. Epidemiol Infect. 2022 Jul 12;150:e143.

40. Dal-Ré R. The winding 12-month journey of the AstraZeneca COVID-19 vaccine since its first administration to humans. Therapie. 2023;78(3):293–302.

41. L’arrivée de 250.000 doses de vaccin Moderna en RDC est une avancée dans la lutte contre la COVID-19 [Internet]. [cited 2023 Oct 30]. Available from: https://www.unicef.org/drcongo/communiques-presse/arrivee-250000-doses-vaccin-moderna-avancee-lutte-covid19

42. Sun J, Uwishema O, Kassem H, Abbass M, Uweis L, Rai A, et al. Ebola virus outbreak returns to the Democratic Republic of Congo: An urgent rising concern. Ann Med Surg. 2022 Jun 6;79:103958.

43. Okonji OC, Rackimuthu S, Gangat SA, Mohanan P, Uday U, Islam Z, et al. Meningitis during COVID -19 pandemic in the Democratic Republic of Congo: A call for concern. Clin Epidemiol Glob Health. 2022;13:100955.

44. Bijukara S, Holland H. Congo virus funds embezzled by ‘mafia network’, says deputy minister. Reuters [Internet]. 2020 Jul 8 [cited 2023 Oct 30]; Available from: https://www.reuters.com/article/health-coronavirus-congo-corruption-idINL4N2EF2H8

45. La Turquie fait don de 100 000 doses de vaccins anti-Covid à la RDC [Internet]. [cited 2023 Oct 30]. Available from: https://www.aa.com.tr/fr/afrique/la-turquie-fait-don-de-100-000-doses-de-vaccins-anti-covid-à-la-rdc/2508096

46. Democratic Republic of the Congo, WHO and UNICEF estimates of immunization coverage: 2021 revision. WHO and UNICEF [Internet]. UNICEF Data. 2022 [cited 2023 Oct 30]. Available from: https://data.unicef.org/wp-content/uploads/2022/07/cod.pdf

47. Amani A, Mossus T, Cheuyem FZL, Bilounga C, Mikamb P, Basseguin Atchou J, et al. Gender and COVID-19 Vaccine Disparities in Cameroon. COVID. 2022;2(12):1715–30.

48. Takougang I, Cheuyem FZL, Lyonga EE, Ndungo JH, Mbopi-Keou FX. Observance of Standard Precautions for Infection Prevention in The Covid-19 Era: A Cross Sectional Study in Six District Hospitals in Yaounde, Cameroon. Am J Biomed Sci Res. 2023;19(5):590–8.

49. Cheuyem FZL, Lyonga EE, Kamga HG, Mbopi-Keou FX, Takougang I. Needlestick and Sharp Injuries and Hepatitis B Vaccination among Healthcare Workers: A Cross Sectional Study in Six District Hospitals in Yaounde (Cameroon). J Community Med Public Health. 2023;7(3):1–9.

